# Saliva is less sensitive than nasopharyngeal swabs for COVID-19 detection in the community setting

**DOI:** 10.1101/2020.05.11.20092338

**Authors:** D. Becker, E. Sandoval, A. Amin, P. De Hoff, A. Diets, N. Leonetti, YW. Lim, C. Elliott, L. Laurent, J. Grzymski, JT Lu

## Abstract

The use of saliva collection for SARS-CoV-2 diagnostics in the ambulatory setting provides several advantages when compared to nasopharyngeal swabs (NPS), including ease of self-collection and reduced use of personal protective equipment (PPE). In addition saliva collection could be advantageous in advising if a convalescent patient is able to return to work after a period of self-quarantine. We investigated the utility of saliva collection in the community setting at Renown Health in a prospective Diagnostic Cohort of 88 patients and in a Convalescent Cohort of 24 patients. In the Diagnostic Cohort, we find that saliva collection has reduced sensitivity (~30% less) relative to NPS. And in our convalescent cohort of patients greater than 8 days and less than 21 days from first symptom, we find that saliva has ~ 50% sensitivity relative to NPS. Our results suggest that rigorous studies in the intended populations should be performed before large-scale screening using saliva as the test matrix is initiated.

## Introduction

Strict activity restrictions have been demonstrated to dramatically reduce the spread of COVID-19 and decrease morbidity and mortality. However, such restrictions have had marked negative economic impacts on communities around the world. It has become clear that accurate broad-based testing for SARS-CoV-2 infection is a critical element in any strategy that will allow communities to safely return to normal activity levels [1,2]. Such testing is important for both detection of new infections, and for determining whether an infected individual is no longer infectious and thus safe to come in contact with uninfected members of the community.

SARS-CoV-2 infection is typically detected by performing real-time reverse transcriptase polymerase chain reaction (RT-PCR) on RNA extracted from samples collected using nasopharyngeal swabs (NPS), although other sample types, such as oropharyngeal swabs and anterior nares swabs, have also been deemed to be acceptable [3]. Swab-based sample collection, particularly NPS, is typically performed by a healthcare provider, is uncomfortable for the patient, and is risky for healthcare providers, as it frequently induces coughing or sneezing. Recent EUA approvals of saliva-based collection have sparked interest in alternative methodologies for safe and simple sample collection [4,5]. This study evaluates the performance of saliva specimens in the community setting for initial diagnosis of SARS-CoV-2 infection in symptomatic patients who meet CDC criteria for clinical testing (Diagnosis Cohort), and also for patients who previously had a positive clinical NPS-based COVID-19 test (Convalescent Cohort) at Renown Health (Reno, NV).

## Methods

This study was reviewed and approved by the University of Nevada, Reno IRB #1582643-6.

### Sample Collection for Diagnosis Cohort

From March 26, 2020 to April 13, 2020, 88 patients underwent simultaneous prospective collection of NPS and saliva specimens for diagnostic evaluation for COVID-19. These patients were identified in a community testing environment. None of these patients were admitted to the hospital at the time of testing. NPS samples were sent for testing at the Nevada State Public Health Testing Laboratory, which performed the CDC COVID-19 assay [6]. Saliva samples were sent via overnight shipping to Helix, a high complexity CLIA laboratory (CLIA #05D2117342, CAP #9382893). 33 and 55 saliva specimens were collected using Orasure OM-505 Microbiome and OGD-610 DNA collection kits, respectively. With both devices, a proprietary nucleic acid stabilization solution is provided with the collection devices and is added by the patient to their saliva immediately after collection. For the first 60 specimens, patients were randomized to either tube. For the subsequent 28 specimens, only the OGD-610 tube was used, as the OM-505 and OGD-610 tubes appeared to have similar performance characteristics for contrived samples. This study was reviewed and improved by the University of Nevada, Reno IRB #1582643-6.

### Sample Collection for Convalescent Cohort

From April 14 to April 24, 2020, 24 patients with prior positive COVID-19 results at Renown Health (Reno, NV) were recalled for subsequent paired NPS and saliva collection. Time since first symptoms and time since the first positive COVID-19 test was captured for clinical correlation. Two of these patients were admitted to the hospital at the time of first testing. Samples collected using NPS in Viral Transport Media (VTM) and saliva samples collected in Orasure OGD-610 DNA collection kits were sent via overnight shipping to Helix. Samples were aliquoted upon receipt and immediately processed. Additional aliquots were stored at −20C for subsequent processing.

### RNA extraction

RNA extractions for NPS VTM and saliva samples (400 uL input volume per sample) were performed at Helix using the MagMax Viral/Pathogen RNA purification kit (ThermoFisher CAT: A42352), with an elution volume of 50 uL.

### RT-PCR assays

RT-PCR assays were set up according to the manufacturers’ protocols with the exception of modifications to the RNA input and reaction volumes, as noted in the main text. RT-PCR assays at Helix were set up manually. RT-PCR assays at the UCSD lab were set up using a Mosquito HV liquid handler (STP Labtech).

### Estimate of Technical Performance with Bayesian Latent Class Models

In scenarios where disease diagnosis is often performed using competing methods, in which neither is a true gold standard, it is possible to estimate test performance (sensitivity, specificity and disease prevalence) using latent class models [7]. Test performance can be parameterized and estimated using bayesian models, where conjugate beta priors can be multiplied with binomial likelihood functions to derive posterior estimates for test performance. We modified an already implemented Gibbs sampler for [8] latent class models in R and used the following hyperparameters (α,β) for the Beta priors for prevalence (α=1, β=4), sensitivity (α=1, β=1) and specificity (α=2.5, β=1) to create weak but reasonable priors.

#### Saliva Collection Devices

The Oragene OGD-610 is a DNA collection kit (https://www.dnagenotek.com/US/products/collection-human/oragene-dx/600-series/QGD-610.html) that provides room temperature and transport stability for DNA. The Oragene OM-505 is a Microbiome Collection Kit (https://www.dnagenotek.com/US/products/collection-microbiome/omnigene-oral/OM-505.html) that provides DNA and RNA stability for up to 3 weeks. The Spectrum S-1000 is a DNA collection device that also provides room temperature and transport stability for DNA (https://spectrumsolution.com/spectrum-dna/clinical-products/sdna-whole-saliva-dna-collection-devices/).

## Results

### Limit of Detection of RT-PCR assays

The Limits of Detection of two sets of RT-PCR assays, the TaqPath Multiplex RT-PCR COVID-19 Kit (Thermo) and the PrimerDesign COVID-19 assay, were determined using different viral RNA and reaction volumes (**Supplementary Table 1**).

The limit of detection of the TaqPath Multiplex RT-PCR COVID-19 Kit (Thermo) was tested in two laboratories using the COVID-19 control RNA from the TaqPath COVID-19 Control Kit (Thermo). In the Helix lab, a miniaturized 5 uL input RNA (10 uL total reaction volume) TaqPath assay was performed on a Quantstudio 7 qRT-PCR instrument (Thermo), and the limit of detection was determined to be 6.25 viral copies. In the UCSD lab, a miniaturized 2 uL input RNA (3 uL total reaction volume) TaqPath assay was performed on a Quantstudio 5 qRT-PCR instrument (Thermo), and the limit of detection was determined to be 3.125 viral copies.

The limit of detection of the PrimerDesign COVID-19 assay was determined at Helix using the Twist SARS-COV-2 synthetic RNA control (Twist Bioscience, Cat. # 102019 and 102024). First, the standard 8ul input RNA (20 uL total reaction volume) PrimerDesign assay was performed using a Roche LightCycler 480 II, and the limit of detection was determined to be 12.5 viral copies (PrimerDesign v1, **Supplementary Table 1**). Next, a miniaturized 2 uL input RNA (5 uL total reaction volume) PrimerDesign assay was performed using a Thermofisher Quantstudio 7, and the limit of detection was determined to be 3.125 viral copies (PrimerDesign v2, **Supplementary Table 1**).

### Detection of SARS-CoV-2 using NPS vs saliva in the Diagnosis Cohort

We compared the performance of saliva-based detection of SARS-CoV-2 to standard NPS-based detection in symptomatic patients who met CDC criteria for standard-of-care clinical testing. A total of 88 individuals who presented and qualified for testing were consented and enrolled in the Diagnosis Cohort. The first 50 subjects in this cohort were randomized to saliva collection using the OM-505 or OGD-610 kits, and the last 18 subjects collected saliva using the OGD-610 kit. NPS samples were sent to the Nevada State Health Lab, which used the CDC RT-qPCR assay for diagnostic testing. Saliva samples were sent to Helix, where RNA was extracted and evaluated using the PrimerDesign COVID-19 assay performed at Helix and theTaqPath Multiplex RT-PCR COVID-19 assay performed at UCSD (**Table 1**).

**Table 1:**
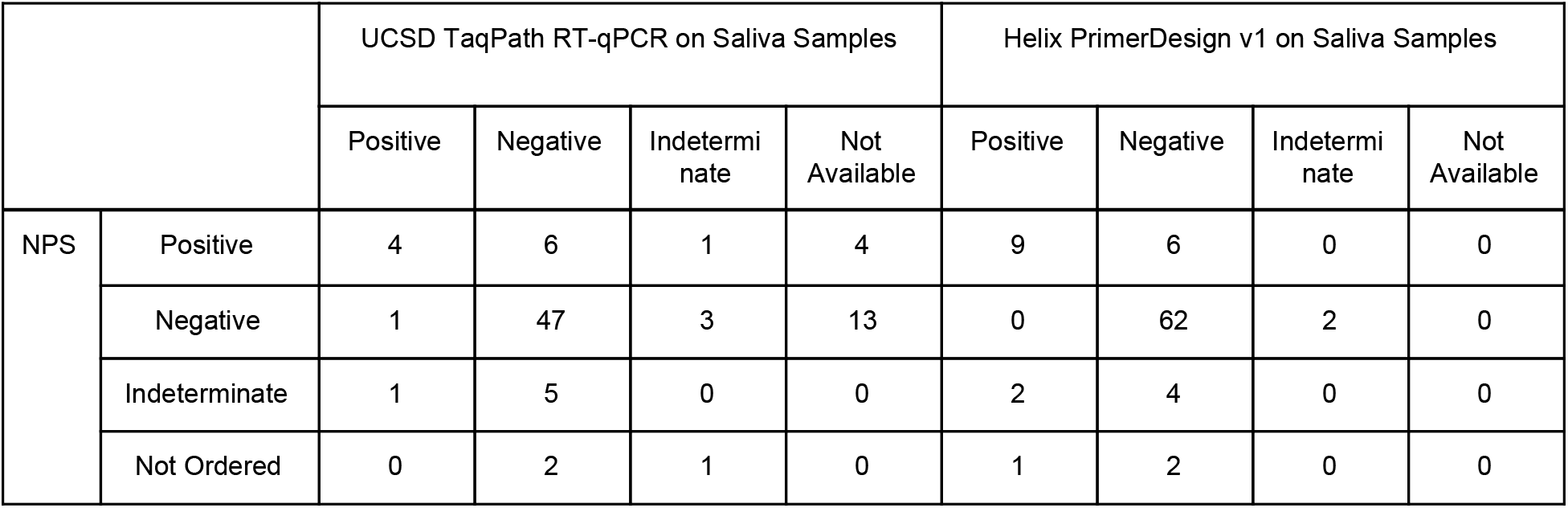
Comparison of qualitative results from NP swabs (NPS) processed at the Nevada State Health Lab and two RT-qPCR assays run on saliva samples collected using OM-505 or OGD-610 saliva collection kits. 3 NPS tests were "Not Ordered” by the physician. Some samples (categorized "Not Available”) were not evaluated at UCSD.

|  | UCSD TaqPath RT-qPCR on Saliva Samples |  |  |  | Helix PrimerDesign v1 on Saliva Samples |  |  |  |
| --- | --- | --- | --- | --- | --- | --- | --- | --- |
|  | Positive | Negative | Indeterminate | Not Available | Positive | Negative | Indeterminate | Not Available |

|  |  |  |  |  |  |  |  |  |  |
| --- | --- | --- | --- | --- | --- | --- | --- | --- | --- |
| NPS | Positive | 4 | 6 | 1 | 4 | 9 | 6 | 0 | 0 |
|  | Negative | 1 | 47 | 3 | 13 | 0 | 62 | 2 | 0 |
|  | Indeterminate | 1 | 5 | 0 | 0 | 2 | 4 | 0 | 0 |
|  | Not Ordered | 0 | 2 | 1 | 0 | 1 | 2 | 0 | 0 |

Using the Helix PrimerDesign v1 results we attempted to estimate the relative performance of saliva to NPS with Bayesian Latent Class Models [7,9]. The NPS sensitivity is estimated to be 98.9% (95% CI: 67.6% -99.7%) and saliva sensitivity is estimated to be 69.2% (95% CI: 38.6% -97.6%), with median reduction in sensitivity of 29.7%.

Subanalysis of the clinical results did not illustrate differences in RNA extraction yield, however Ct values for the internal extraction control (where available) and for samples with positive Ct values for Orf1ab (RdRp) were lower on OGD-610 (**Supplementary Figure 1**). Based on these findings, we decided to use only the OGD-610 device for saliva collection in the Convalescent Cohort.

### Detection of SARS-CoV-2 using NPS vs saliva in the Convalescent Cohort

To compare the performance of saliva-based testing to standard NPS-based testing in convalescent cases, Renown recalled known COVID-19 positive cases for paired collection. RNA was extracted from the NPS VTM and Saliva samples and analyzed using the PrimerDesign and TaqPath assays at the sites shown in **Table 2** (full dataset with Ct values for each viral target sequence and internal control sequences are shown in **Supplementary Table 2**). Concordance in positive and negative results for the TaqPath assay between the two sites, UCSD and Helix, was excellent. There was only one sample, the NPS VTM sample, from Subject 11, which was positive for the TaqPath assay at one site (UCSD) and negative at the other site (Helix). The two assays, TaqPath and PrimerDesign, were discordant for four samples, with the PrimerDesign assay being generally less sensitive: the NPS VTM samples were negative using the PrimerDesign assay and positive for the TaqPath assay for Subjects 13, 18, and 8; and for the NPS VTM sample for Subject 11, the Helix PrimerDesign and TaqPath assays were negative, but the UCSD TaqPath assay was positive.

**Table 2.** Summary of Results from Convalescent Cohort. *”Date since first positive test” as “Date since first symptom” was not known.

| Paired Subject ID | Days since first symptom | Helix PrimerDesign |  | UCSD TaqPath |  | Helix TaqPath |  |
| --- | --- | --- | --- | --- | --- | --- | --- |
|  |  | NPS VTM | Saliva | NPS VTM | Saliva | NPS VTM | Saliva |
| 19 | 8 | POSITIVE | POSITIVE | POSITIVE | POSITIVE | POSITIVE | POSITIVE |
| 32 | 9 | POSITIVE | ND | *Not Tested* | *Not Tested* | POSITIVE | POSITIVE |
| 6 | 11 | ND | ND | ND | INCONCLUSIVE | ND | INCONCLUSIVE |
| 7 | 11 | ND | ND | INCONCLUSIVE | ND | ND | ND |
| 20 | 12 | POSITIVE | POSITIVE | *Not Tested* | *Not Tested* | POSITIVE | POSITIVE |
| 30 | 12 | POSITIVE | ND | *Not Tested* | *Not Tested* | POSITIVE | ND |
| 23 | 12 | ND | ND | *Not Tested* | *Not Tested* | *Not Tested* | *Not Tested* |
| 1 | 13 | ND | ND | INCONCLUSIVE | INCONCLUSIVE | ND | ND |
| 13 | 14 | ND | ND | POSITIVE | ND | POSITIVE | ND |
| 14 | 14 | ND | ND | INCONCLUSIVE | ND | INCONCLUSIVE | ND |
| 33 | 14 | POSITIVE | ND | *Not Tested* | *Not Tested* | POSITIVE | ND |
| 18 | 15 | ND | ND | POSITIVE | INCONCLUSIVE | POSITIVE | INCONCLUSIVE |
| 17 | 16 | ND | ND | ND | ND | *Not Tested* | *Not Tested* |
| 8 | 18 | INVALID | ND | POSITIVE | ND | POSITIVE | ND |
| 21 | 18 | POSITIVE | POSITIVE | *Not Tested* | *Not Tested* | POSITIVE | POSITIVE |
| 10 | 24 | ND | POSITIVE | ND | POSITIVE | ND | POSITIVE |
| 16 | 25* | ND | ND | ND | ND | *Not Tested* | *Not Tested* |
| 22 | 27 | ND | INVALID | *Not Tested* | *Not Tested* | *Not Tested* | *Not Tested* |
| 9 | 28 | ND | ND | ND | ND | *Not Tested* | *Not Tested* |
| 3 | 30 | ND | ND | INCONCLUSIVE | ND | INCONCLUSIVE | ND |
| 15 | 35 | ND | ND | ND | ND | *Not Tested* | *Not Tested* |
| 4 | 37 | ND | ND | ND | ND | *Not Tested* | *Not Tested* |
| 2 | 42 | ND | ND | ND | ND | *Not Tested* | *Not Tested* |
| 12 | 56 | ND | ND | ND | ND | *Not Tested* | *Not Tested* |
| 5 | n/a | ND | ND | ND | ND | *Not Tested* | *Not Tested* |
| 11 | n/a | ND | ND | POSITIVE | ND | ND | ND |

Overall, the sensitivity using the NPS VTM samples was higher than for the Saliva samples. Looking at the results from the more sensitive Helix TaqPath assay, both sample types yielded positive results for 4 Subjects, the NPS VTM only was positive for 5 Subjects, and the Saliva only was positive for 1 Subject. As expected, the positive rate decreased for both sample types over time. Of the 15 Subjects who were tested using the Helix TaqPath assay within three weeks of their first symptom, 9 had positive results from the NPS VTM sample and 4 had positive results from the Saliva sample. For the 9 Subjects who were tested more than three weeks after their first symptom, none had a positive result from the NPS VTM and only 1 had a positive result from the Saliva sample. We note that several Subjects had Inconclusive results for one or both samples; this call results from one out of the three viral target sequences yielding a detectable results for the TaqPath assay and may reflect low viral titer; in nearly every case, the positive target was the N gene, which usually yields the lowest Ct value of the three targets. There were 2 subjects for which the number of days from the first symptom was unknown or could not be corroborated.

## Discussion

Self-collected saliva for SARS-CoV-2 testing could reduce patient discomfort and risk of viral transmission to healthcare workers, and also alleviate supply shortages for NPS and viral transport media. Saliva collection has been reported to perform as well or better than NPS for detection of SARS-CoV-2 infection [4,5]. The results of this study do not corroborate these previously reported findings.

We believe there are several plausible explanations for these different outcomes. First, our study focused on enrolling subjects from the community who were diagnosed as outpatients; only 2 patients in the entire study were inpatients, and neither was admitted to the ICU. Prior studies largely focused on inpatient populations, of which a large proportion were ICU patients. Outpatients are likely to have milder symptoms, and have been shown to have reduced viral titers relative to more acute patients [10,11]. Our findings are lower than a large community study of 622 patients with paired NPS and saliva samples, where saliva samples were used to confirm NPS findings. In this study, only 84.9% of NPS samples were confirmed with saliva. This reduced sensitivity seems to be related to reduced viral titer in saliva samples [12] as well as differences in temporal dynamics in shedding in upper respiratory locations versus saliva [13,14]. Lastly, in convalescent patients, viral titers remain at detectable levels up to 21 days after initial testing [15,16]. Unfortunately, in this population, our saliva results had only ~50% of the sensitivity of NPS.

All reported studies have used different saliva collection methodologies. Different preservation solutions may differ in their ability to protect viral RNA from degradation, and can also interact with extraction chemistry, potentially impacting the efficiency of RNA extraction or causing inhibition of the qPCR reaction. To further evaluate whether the particular preservation solution we used may have impacted the results, we evaluated side by side, VTM, the OGD-610 and the Spectrum S-1000, which was used in one of the previous studies [4]. The samples were prepared with a negative clinical matrix spiked with synthetic viral RNA sequences at 4000, 1000, 500 and 200 GCE/ ml. We did not observe decreased extraction efficiencies or inhibition between the OGD-610, which was used in our study, and the Spectrum S-1000 (**Supplementary Table 3**).

The limitations of this study include the lack of detailed clinical information about participating patients; we did not attempt to correlate clinical characteristics with test outcomes. Further investigation and confirmation of this study is warranted. We acknowledge that alternative saliva collection methods with different preservation solutions, different extraction chemistries, or use of a more sensitive COVID-19 assay may yield better results in mildly symptomatic patients in the community setting. We are hopeful that further studies of these variables will provide an alternative collection system for testing mildly symptomatic or convalescent patients in the community setting. Our results suggest that rigorous studies in the intended populations should be performed before large-scale screening using saliva as the test matrix is initiated.

## Data Availability

The datasets generated during and/or analyzed during the current study are available from the corresponding author on reasonable request.

## Supplementary Materials

**Supplementary Table 1:** **Limit of Detection for direct viral copies, using Twist RNA or COVID-19 control RNA from the TaqPath COVID-19 Control Kit, into RT-qPCR for Helix PrimerDesign and TaqPath assays, and for the UCSD Taqpath assay. Values represent Mean Cq values., using a Cq confidence cut off of >0.8. ND= Not Determined**.

|  | Helix |  |  | UCSD |
| --- | --- | --- | --- | --- |
| Copies | PrimerDesign v1 | PrimerDesign v2 | TaqPath | TaqPath |
| Target | ORF1ab | ORF1ab | S, N, ORF1ab | S, N, ORF1ab |
| 100 | 35.85 | 31.56 | S: 30.29 | S: 27.99 |
|  |  |  | N: 31.66 | N: 27.26 |
|  |  |  | ORF1ab: 30.19 | ORF1ab: 28.02 |
| 50 | 36.11 | 32.43 | 31.38 | 28.51 |
|  |  |  | 32.62 | 28.90 |
|  |  |  | 31.20 | 28.73 |
| 25 | 36.75 | 33.90 | 31.42 | 29.43 |
|  |  |  | 33.19 | 29.67 |
|  |  |  | 31.90 | 29.65 |
| 12.5 | 37.85 | 33.75 | 32.09 | 30.74 |
|  |  |  | 33.25 | 29.88 |
|  |  |  | 32.10 | 29.98 |
| 6.25 | ND | 35.38 | 32.29 | 31.33 |
|  |  |  | 34.31 | 31.45 |
|  |  |  | 32.52 | 31.24 |
| 3.125 | ND | 36.67 | ND | 35.21 |
|  |  |  | 33.20 | 33.02 |
|  |  |  | ND | 34.06 |

**Supplementary Figure 1:**
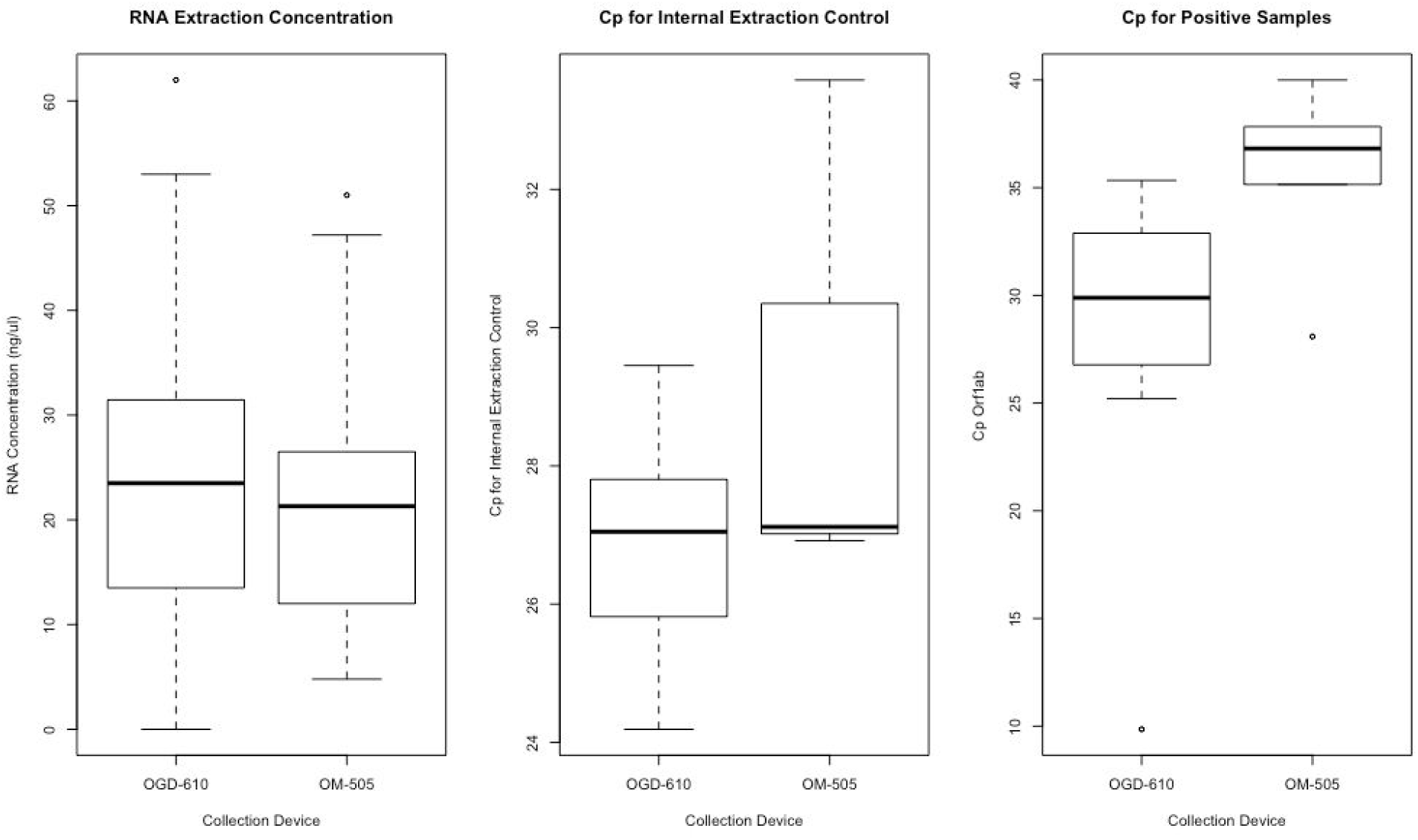
**(a) Comparison of RNA Extraction Yield where available for OGD-610 vs OM-505, (b) Comparison of Cp value for PrimerDesign Internal Extraction Control and (c) Comparison of Cp value for PrimerDesign Internal Extraction (for a subset of samples only), Control Cp and viral gene target Cpt and viral gene target Ct on OGD-610 and OM-505 on positive clinical samples from Diagnosis Cohort**

**Supplementary Table 2:** **For Convalescent Cohort, Cp values for each viral target sequence and internal extraction control sequences for Primer Design. For TaqPath, MS2 phage was not spiked into the samples at the time of extraction of these clinical samples. Values represent Mean Cp values., using a Cq confidence cut off of >0.8. ND= Not Determined**.

**2a. Samples with SARS-CoV-2 is detectable in at least one specimen type**
| ID # | PrimerDesign v2 |  |  |  | Helix TaqPath |  |  |
| --- | --- | --- | --- | --- | --- | --- | --- |
|  | Target | VTM Cp | OGD-610 Cp |  | Target | VTM Cp | OGD-610 Cp |
| 8 | RdRp | ND | ND |  | N gene | 33.99 | ND |
|  | PD-IEC | ND | 21.86 |  | ORF1ab | 31.44 | ND |
|  |  |  |  |  | S gene | ND | ND |
| 10 | RdRp | ND | 37.12 |  | N gene | ND | 29.43 |
|  | PD-IEC | 30.10 | 25.09 |  | ORF1ab | ND | 30.35 |
|  |  |  |  |  | S gene | ND | 31.46 |
| 13 | RdRp | ND | ND |  | N gene | 33.57 | ND |
|  | PD-IEC | 29.64 | 24.68 |  | ORF1ab | ND | ND |
|  |  |  |  |  | S gene | 33.19 | ND |
| 18 | RdRp | ND | ND |  | N gene | 32.45 | 33.47 (LOW Q) |
|  | PD-IEC | 34.62 | 24.79 |  | ORF1ab | 30.79 | ND |
|  |  |  |  |  | S gene | 30.73 | ND |
| 19 | RdRp | 34.73 | 37.57 |  | N gene | 31.18 | 30.56 |
|  | PD-IEC | 31.04 | 22.79 |  | ORF1ab | 29.77 | 32 |
|  |  |  |  |  | S gene | 29.56 | ND |
| 20 | RdRp | 26.95 | 35.30 |  | N gene | 20.35 | 31.72 |
|  | PD-IEC | 40.09 | 23.45 |  | ORF1ab | 21.14 | 31.37 |
|  |  |  |  |  | S gene | 21.72 | 31.39 |
| 21 | RdRp | 22.05 | 38.19 |  | N gene | 19.03 | 31.17 |
|  | PD-IEC | ND | 25.29 |  | ORF1ab | 18.09 | ND |
|  |  |  |  |  | S gene | 18.67 | ND |
| 30 | RdRp | 29.92 | ND |  | N gene | 26.62 | ND |
|  | PD-IEC | ND | 23.89 |  | ORF1ab | 25.77 | ND |
|  |  |  |  |  | S gene | 26.17 | ND |
| 32 | RdRp | 32.93 | ND |  | N gene | 28.87 | 28.87 |
|  | PD-IEC | 23.95 | 26.41 |  | ORF1ab | 28.40 | 28.4 |
|  |  |  |  |  | S gene | 28.73 | 28.73 |
| 33 | RdRp | 31.11 | ND |  | N gene | 27.81 | ND |
|  | PD-IEC | 23.74 | 25.26 |  | ORF1ab | 27.40 | ND |
|  |  |  |  |  | S gene | 27.81 | ND |

2b. Samples where SARS-CoV-2 was not detectable in either specimen type
| ID # | PrimerDesign |  |  |
| --- | --- | --- | --- |
|  | Target | VTM Cp | OGD-610 Cp |
| 1 | RdRp | ND | ND |
|  | PD-IEC | 31.80 | 22.55 |
| 2 | RdRp | ND | ND |
|  | PD-IEC | 34.84 | 23.52 |
| 3 | RdRp | ND | ND |
|  | PD-IEC | 35.06 | 22.30 |
| 4 | RdRp | ND | ND |
|  | PD-IEC | 34.03 | 22.74 |
| 5 | RdRp | ND | ND |
|  | PD-IEC | 32.69 | 26.76 |
| 6 | RdRp | ND | ND |
|  | PD-IEC | 32.45 | 24.79 |
| 7 | RdRp | ND | ND |
|  | PD-IEC | 29.79 | 22.77 |
| 9 | RdRp | ND | ND |
|  | PD-IEC | 31.95 | 24.78 |
| 11 | RdRp | ND | ND |
|  | PD-IEC | 32.44 | 24.56 |
| 12 | RdRp | ND | ND |
|  | PD-IEC | 33.27 | 23.24 |
| 14 | RdRp | ND | ND |
|  | PD-IEC | 32.05 | 25.20 |
| 15 | RdRp | ND | ND |
|  | PD-IEC | 30.41 | 23.67 |
| 16 | RdRp | ND | ND |
|  | PD-IEC | 27.74 | 22.73 |
| 17 | RdRp | ND | ND |
|  | PD-IEC | 29.75 | 26.20 |
| 22 | RdRp | ND | ND |
|  | PD-IEC | 34.95 | ND |
| 23 | RdRp | ND | ND |
|  | PD-IEC | 31.73 | 25.54 |

**Supplementary Table 3:** Limit of Detection comparison of VTM, OGD-610, and Spectrum S-1000 in Contrived Samples. The Spectrum S-1000 saliva collection kit was used in a previous study that showed good correlation between NPS and saliva specimens. We evaluated the performance of both saliva collection tubes side by side, in comparison with VTM using Twist RNA control spike-in prior. Values represent Mean Cp values, using a Cq confidence cut off of >0.8. ND= Not Determined.

|  | Helix TaqPath |  |  |  |
| --- | --- | --- | --- | --- |
| Copies | Target | VTM | OGD-610 | Spectrum S-1000 |
| 4000/ml | S Gene | 30.63 | 30.89 | 31.09 |
|  | N Gene | 33.93 | 31.99 | 32.86 |
|  | ORF1ab | 30.87 | 31.85 | 32.29 |
| 1000/ml | S Gene | 32.37 | ND | ND |
|  | N Gene | 34.67 | 32.17 | ND |
|  | ORF1ab | 32.86 | 33.29 | ND |
| 500/ml | S Gene | 32.42 | ND | ND |
|  | N Gene | 34.94 | ND | ND |
|  | ORF1ab | 33.39 | 32.69 | ND |
| 200/ml | S Gene | ND | ND | ND |
|  | N Gene | 33.67 | ND | ND |
|  | ORF1ab | ND | ND | ND |

